# Measurement Matters: Changing Penalty Calculations under the Hospital Acquired Condition Reduction Program (HACRP) Cost Hospitals Millions

**DOI:** 10.1101/2020.10.19.20215194

**Authors:** Olga A. Vsevolozhskaya, Karina C. Manz, Pierre M. Zephyr, Teresa M. Waters

## Abstract

**Background:** Since October 2014, the Centers for Medicare and Medicaid Services has penalized 25% of U.S. hospitals with the highest rates of hospital-acquired conditions under the Hospital Acquired Conditions Reduction Program (HACRP). While early evaluations of the HACRP program reported cumulative reductions in hospital-acquired conditions, more recent studies have not found a clear association between receipt of the HACRP penalty and hospital quality of care. We posit that some of this disconnect may be driven by frequent scoring updates. The sensitivity of the HACRP penalties to updates in the program’s scoring methodology has not been independently evaluated.

**Methods:** We used hospital discharge records from 14 states to evaluate the association between changes in HACRP scoring methodology and corresponding shifts in penalty status. To isolate the impact of changes in scoring methods over time, we used FY2018 hospital performance data to calculate total HAC scores using FY2015 through FY2018 CMS scoring methodologies.

**Results:** Comparing hospital penalty status based on various HACRP scoring methodologies over time, we found a significant overlap between penalized hospitals when using FY 2015 and 2016 scoring methodologies (95%) and between FY 2017 and 2018 methodologies (46%), but substantial differences across early vs later years. Only 15% of hospitals were eligible for penalties across all four years. We also found significant changes in a hospital’s (relative) ranking across the various years, indicating that shifts in penalty status were not driven by small changes in HAC scores clustered around the penalty threshold.

**Conclusions:** HACRP penalties have been highly sensitive to program updates, which are generally announced after performance periods are concluded. This disconnect between performance and penalties calls into question the ability of the HACRP to improve patient safety as intended.

## Background

Hospital-acquired conditions (HACs) place patients at higher risk for future health complications and may indicate poor hospital quality of care. Since October 2014, the Centers for Medicare and Medicaid Services (CMS), under the Hospital-Acquired Condition Reduction Program (HACRP), has penalized hospitals in the worst performing quartile of HAC quality measures; that is, hospitals with a total HAC score above the 75th percentile are subject to a one percent penalty on their Medicare revenues, assessed when CMS pays a claim.

Penalties assessed under HACRP are sizeable (Fig. 1). While early evaluations of HACRP reported cumulative reductions in hospital-acquired conditions [1, 2], more recent studies [3, 4, 5, 6, 7] have not found a clear association between receipt of the HACRP penalty and hospital quality of care. For example, recent evidence suggests that HACRP penalized hospitals had more accreditations for quality, offered a larger number of advanced services, were major teaching institutions, and had better performance on other process and outcome measures [7]. Other research found that under HACRP, CMS assigned different scores to hospitals whose performance was statistically the same and penalized 25% of hospitals regardless of the statistical significance of the difference between their performance and others [4].

**Figure 1.**
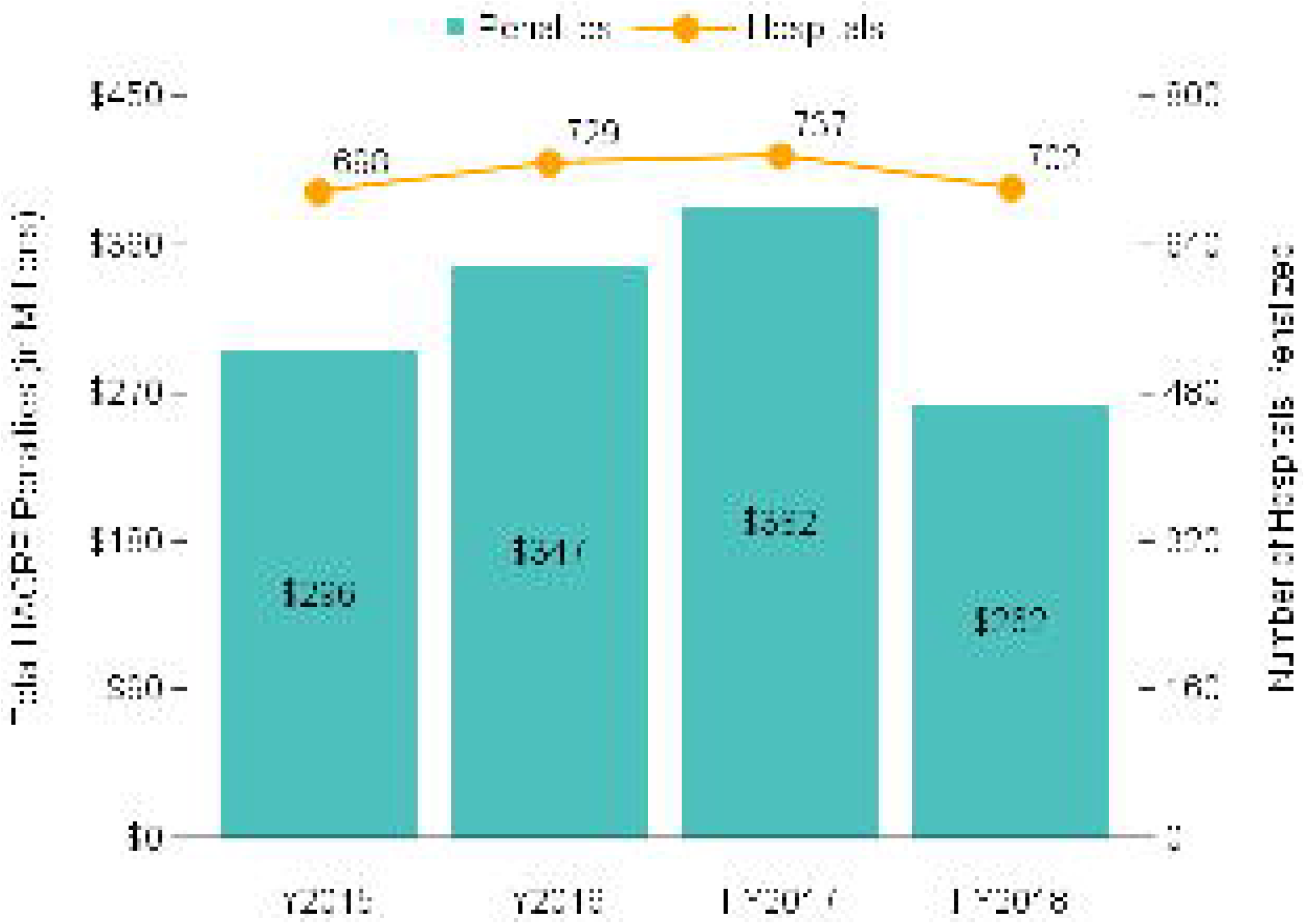
Total HACRP penalties (in millions) by fiscal year based on RAND hospital data [24] merged with total HAC scores, FYs 2015-2018

The empirical link between HACRP penalties and hospital quality of care may be further dampened by repeated changes to the program’s scoring methodology. The total HAC score is a weighted sum of two domain scores. Domain 1 is based on the Agency for Healthcare Research and Quality (AHRQ) Patient Safety Indicator 90 (PSI-90) composite score [8]. Domain 2 is based on the Centers for Disease Control and Prevention (CDC) National Healthcare Safety Network (NHSN) Healthcare Associate Infection (HAI) measures. Both Domain 1 and Domain 2 measures, as well as their individual weights in the total HAC score, undergo annual updates to address stakeholder concerns and better align measures with policy goals.

As one of Medicare’s largest hospital pay-for-performance programs, HACRP is intended to support the CMS goal of linking Medicare payments to inpatient quality of care. Sensitivity of the HACRP penalties to annual program updates, rather than differences in hospital performance, is especially salient given Medicare’s planned expansion of value-based payment models. Using Medicare hospital discharge and NHSN data for hospitals in 14 states, and HACRP scoring methodologies for fiscal years (FYs) 2015-2018, we examined the impact of the changing HACRP scoring algorithms on hospital penalty status.

## Methods

### Data Source and Study Design

Several data sources were used for this hospital-level analysis.

First, we identified Medicare discharges in Healthcare Cost and Utilization Project State Inpatient Databases [9] (HCUP SIDs) for 14 states (Arkansas, Arizona, California, Colorado, Florida, Idaho, Kentucky, Massachusetts, North Caroline, Nebraska, New Jersey, New York, Oregon, and Washington). These states were selected because their discharge data contained sufficient identifying information to be connected with other hospital-level data, and they offered sufficient volume and geographic coverage to provide meaningful insights on the impact of the HACRP scoring methods. Second, we used CDC NHSN standardized infection ratios (SIRs), hospital-level observed-to-predicted numbers of healthcare associated infections (HAIs), available through the Hospital Compare Data Archive [10]. These data files included SIRs for Central Line-Associated Bloodstream Infection (CLABSI), Catheter-Associated Urinary Tract Infection (CAUTI), Surgical Site Infection (SSI), Methicillin-resistant Staphylococcus aureus (MRSA) bacteremia, and Clostridium difficile Infection (CDI). We merged the HCUP SID measures and SIR rates with hospital-level data available through the American Hospital Association Annual Survey of Hospitals.

Our objective was to use one set of hospital performance data and compare the assignment of penalties across four years of the HACRP scoring methodologies. To that end, we selected data for HACRP’s FY2018 performance evaluation period: July 2014 through September 2015 (15 months) of HCUP SID administrative data and January 2015 through December 2016 (24 months) of NHSN HAIs. Then, to isolate the impact of changes in scoring methods over time, we used these FY2018 data to calculate total HAC scores using FY2015 through FY2018 CMS scoring methodologies. We settled on this “retrospective” comparison for several reasons. First, the FY2018 performance evaluation period was the last one to use International Statistical Classification of Diseases (ICD)-9 data before the switch to ICD-10. Second, FY2018 data had enhanced present on admission (POA) reporting compared to earlier performance evaluation periods [11].

### Domain 1 Measure Changes

The AHRQ PSI-90 measure, used as the HACRP Domain 1 score, was designed to provide a simple and transparent metric to understand, communicate, and track patient safety across different US hospitals [12]. PSI-90 is a weighted composite of selected component Patient Safety Indicators (PSIs) and calculated based on hospital administrative data (claims or discharge records). Weights assigned to the individual PSIs that comprise PSI-90 have changed significantly over time (Table 1).

**Table 1.** The AHRQ PSI-90 component measures, by version and year(s), with corresponding weights.

| AHRQ PSI-90 software version | PSI-90 individual weights/POA |  | Recalibrated PSI-90 weights |
| --- | --- | --- | --- |
|  | 4.5a (FY15-16) | 5.0.1 (FY17) | 6.0.2 (FY18) |
| Pressure Ulcer (PSI-03) | 0.2303/0.0220 | 0.0391 | 0.0557 |
| Iatrogenic Pneumothorax (PSI-06) | 0.0545/0.0708 | 0.0905 | 0.0493 |
| Central Line-associated Bloodstream Infections (PSI-07) | 0.0563/0.0652 | 0.0301 | – |
| Postoperative Hip Fracture (PSI-08) | 0.0025/0.0011 | 0.0025 | 0.0093 |
| Perioperative Hemorrhage and Hematoma (PSI-09) | – | – | 0.0820 |
| Postoperative Acute Kidney Injury (PSI-10) | – | – | 0.0466 |
| Postoperative Respiratory Failure (PSI-11) | – | – | 0.3109 |
| Perioperative Pulmonary Embolism or Deep Vein Thrombosis (PSI-12) | 0.2572/0.2579 | 0.3570 | 0.1954 |
| Postoperative Sepsis (PSI-13) | 0.0603/0.0742 | 0.0798 | 0.2318 |
| Postoperative Wound Dehiscence (PSI-14) | 0.0097/0.0165 | 0.0183 | 0.0121 |
| Accidental Puncture or Laceration (PSI-15) | 0.3292/0.4917 | 0.3827 | 0.0067 |

POA is Present on Admission indicator; if POA available in the data a different set of weights was used by the AHRQ software.

In FYs 2015 and 2016, CMS calculated hospital-level PSI-90 measures based on the AHRQ PSI software version 4.5a [13]. Rates of individual hospital-acquired conditions were weighted based on their volume, using 2012 HCUP SID all-payer discharge data for patients 18 and older [12].This weighting approach did not necessarily reflect corresponding harm. For example, PSI-15 was weighted heavily because it occurred frequently; however, a minor puncture may not have resulted in severe harm. Weighting based on volume provided hospitals an opportunity to improve their performance by targeting frequently occurring PSIs rather than the most harmful [14, 15].

In FY 2017, PSI-90 component measure weights were reviewed and a reweighted composite measure was calculated by CMS using the AHRQ PSI software version [16]. The recalibrated software modified PSI-90 composite weights based on component PSI volumes derived from the July 2012 - June 2014 Medicare FFS claims data, rather than 2012 HCUP SID all-payer discharge data for patients 18 and older (as in AHRQ PSI version 4.5a).

In FY 2018, in response to the concerns that the PSI-90 weighting scheme was based on the volume of PSIs rather than associated harm, CMS used a new set of weights (incorporated in the AHRQ PSI software version 6.0.2) to calculate the HACRP Domain 1 score [17]. These new weights incorporated volume and harm of individual PSIs according to a severity index. Additionally, PSI-07 (central line-associated bloodstream infections) was removed to avoid double counting of CLAB-SIs as a component of PSI-90 under Domain 1 and as a separate outcome under Domain 2.

### Domain 2 Measure Changes

The National Healthcare Safety Network (NHSN) healthcare-associated infections (HAIs) data have been used to construct HACRP Domain 2 scores.

In the first year of the HACRP (FY 2015) [13], Domain 2 included SIRs for Central Line-Associated Bloodstream Infections (CLABSI) and Catheter-Associated Urinary Tract Infection (CAUTI). Domain 2 scores were expanded to include Surgical Site Infection (SSI) in FY 2016, followed by Methicillin-resistant Staphylococcus aureus (MRSA) bacteremia and Clostridium difficile Infection (CDIF) in FYs 2017 and 2018 [17]. In addition, in FY 2018 Domain 2 CLABSI and CAUTI SIRs were expanded from ICU only measures to include both ICU and ward patients.

### Changes in HACRP Scoring Mythology

To determine a hospital’s total HAC Score, CMS used a decile-based scoring methodology in FY 2015, 2016, and 2017, but switched to the winsorized *z*-score methodology in FY 2018.

Under the decile-based scoring methodology, each hospital received two relative ranking scores, ranging from 1 to 10, based on their Domain 1 and Domain 2 measures. A hospital’s Domain 1 score was determined solely by their PSI-90, while their Domain 2 score was determined by reported HAI measures. Hospitals with at least one HAI score reported to NHSN received a Domain 2 score based on the average of their reported SIRs. Hospitals without at least one reported HAI score either (a) did not receive a Domain 2 score because they had a HAI exception, and their Total HAC score was based solely on Domain 1 performance or (b) had the Domain 2 score set to the maximum (i.e., 10 points) [13, 16].

Unlike the decile-based approach, the winsorized *z*-score methodology uses continuous scores based on raw measures (instead of the relative rank from 1 to 10). If a hospital’s individual HAI measure falls below the 5th percentile (or above the 95th percentile), based on all eligible hospitals reporting the measure, it is set equal to the 5th (95th) percentile value. Then, each HAI measure is converted into a standardized *z*-score. If a hospital did not submit any Domain 2 measures and did not receive a HAI exception, the maximum winsorized *z*-score for Domain 2 was applied; otherwise, no Domain 2 score is calculated.

For FY 2015, CMS applied a weight of 35% for Domain 1 and 65% for Domain 2 to determine the total HAC score; for FY 2016, this changed to 25% for Domain 1 and 75% for Domain 2. In FY 2017 and 2018 the weights changed again: 15% for Domain 1 and 85% for Domain 2. As noted above, if a hospital only had data for one domain score, CMS applied 100% weight to this domain. Under the decile-based scoring approach (FY 2015-17), a hospital’s Total HAC score represented the decile of a hospital’s performance, ranging between 1 and 10. Under the winsorized z-score method (FY 18), total HAC scores ranged from *−*3 to +3. In all years, hospitals with a total HAC score above the 75th percentile of the distribution were subject to a 1% payment reduction.

### Analytic Approach

Total HAC scores for FYs 2015-2018 served as our main study outcome [18] (Table 2).

**Table 2.** Measures adopted for Hospital-Acquired Conditions Reduction Program by fiscal year.

|  | Measure | FY 2015 | FY 2016 | FY 2017 | FY 2018 |
| --- | --- | --- | --- | --- | --- |
| Domain 1 | PSI-90 composite claims-based measure | ✓ | ✓ | ✓ | – |
|  | Recalibrated PSI-90 composite claims-based measure | – | – | – | ✓ |
| Domain 2 | CLABSI | ✓ | ✓ | ✓ | ✓ |
|  | CAUTI | ✓ | ✓ | ✓ | ✓ |
|  | SSI-colon and hysterectomy | – | ✓ | ✓ | ✓ |
|  | MRSA bacteremia | – | – | ✓ | ✓ |
|  | CDI | – | – | ✓ | ✓ |
| Weights |  | Dom1: 65% | Dom1: 75% | Dom1: 85% | Dom1: 85% |
|  |  | Dom2: 35% | Dom2: 25% | Dom2: 15% | Dom2: 15% |

We identified *n* = 1704 hospitals in 14 states for which Domain 1 scores could be calculated and *n* = 1105 hospitals in those states for which at least one Domain 2 measure was reported to NHSN for the evaluation period of interest. For each hospital, the total HAC score was calculated based on FY 2015, 2016, 2017, and 2018 scoring methodologies in two different ways. First, since we did not have information whether a hospital received the HAI exception, we assumed that hospitals without a Domain 2 measure had the required exception and the total HAC score was equal to the Domain 1 value. In a second (alternative) analysis, we assumed that hospitals without a Domain 2 measure did not have the required exception and maximum Domain 2 score was employed. Then, a binary variable (1 = yes or 0 = no) of whether a hospital received a payment penalty under a specific scoring methodology was assigned based on a hospital’s percentile score within the study sample. This information was used to assess the degree to which changes in HACRP scoring algorithms affected hospital penalty status.

The changing scope of CLABSI and CAUTI measures employed by HACRP over the study time period (expanded from ICUs only to ICUs plus select wards in FY 2018 scoring) also required us to predict certain ICU-only measures not included in data publicly available through the Hospital Compare Data Archive. To construct Domain 2 scores under FYs 2015-2017 methodologies, we used an extreme gradient boosting technique, implemented through R xgboost software package [19], to predict FY 2016 CLABSI and CAUTI SIRs for ICUs only. We performed a 70/30 split of the FY 2015 data to train and evaluate a predictive model. Hospital-level predictor variables included: combined SIRs for ICUs and selected wards for FY 2016, total beds, admissions, FTE personnel, and percent of inpatient days covered by Medicare and Medicaid. Root mean square error (RMSE) was used to evaluate prediction accuracy.

## Results

Total HAC scores and their summary characteristics are provided in Additional files 1: Table A1.

A total of 1704 hospitals were assigned a total HAC score for each fiscal year, with 426 above the 75th percentile cutoff value for penalty assessment. As noted above, the empirical 75th percentiles for our data did not exactly matched the CMS-reported cutoffs (Additional files 1: Table A1). Nonetheless, total HAC score cutoffs that were calculated under the assumption that no data submitted to NHSN implied no Domain 2 score closely matches CMS-reported cutoff values, while the alternative approach significantly exceeded these cutoffs. For this reason, our main results, reflecting the assumption “no data submitted to NHSN implied no Domain 2 score,” are presented below, while our alternative results reflecting the assumption “no data submitted to NHSN implies maximum Domain 2 score” are presented in Additional files 1. Because the two approaches simply shift total HAC scores (by adding the maximum Domain 2 value versus zero), relative rankings are not significantly changed, and the two sets of results support the same conclusions.

Comparing hospital penalty status based on various HACRP scoring methodologies over time, we found a significant overlap between penalized hospitals when using FY 2015 and 2016 scoring methodologies (95%) and between FY 2017 and 2018 methodologies (46%), but substantial differences across early vs later years (Fig. 2; Additional files 1 Fig. A3 is a version of this figure but with maximum Domain 2 score values assigned). In addition, only 15% of hospitals were eligible for penalties across all four years/scoring methodologies.

**Figure 2.**
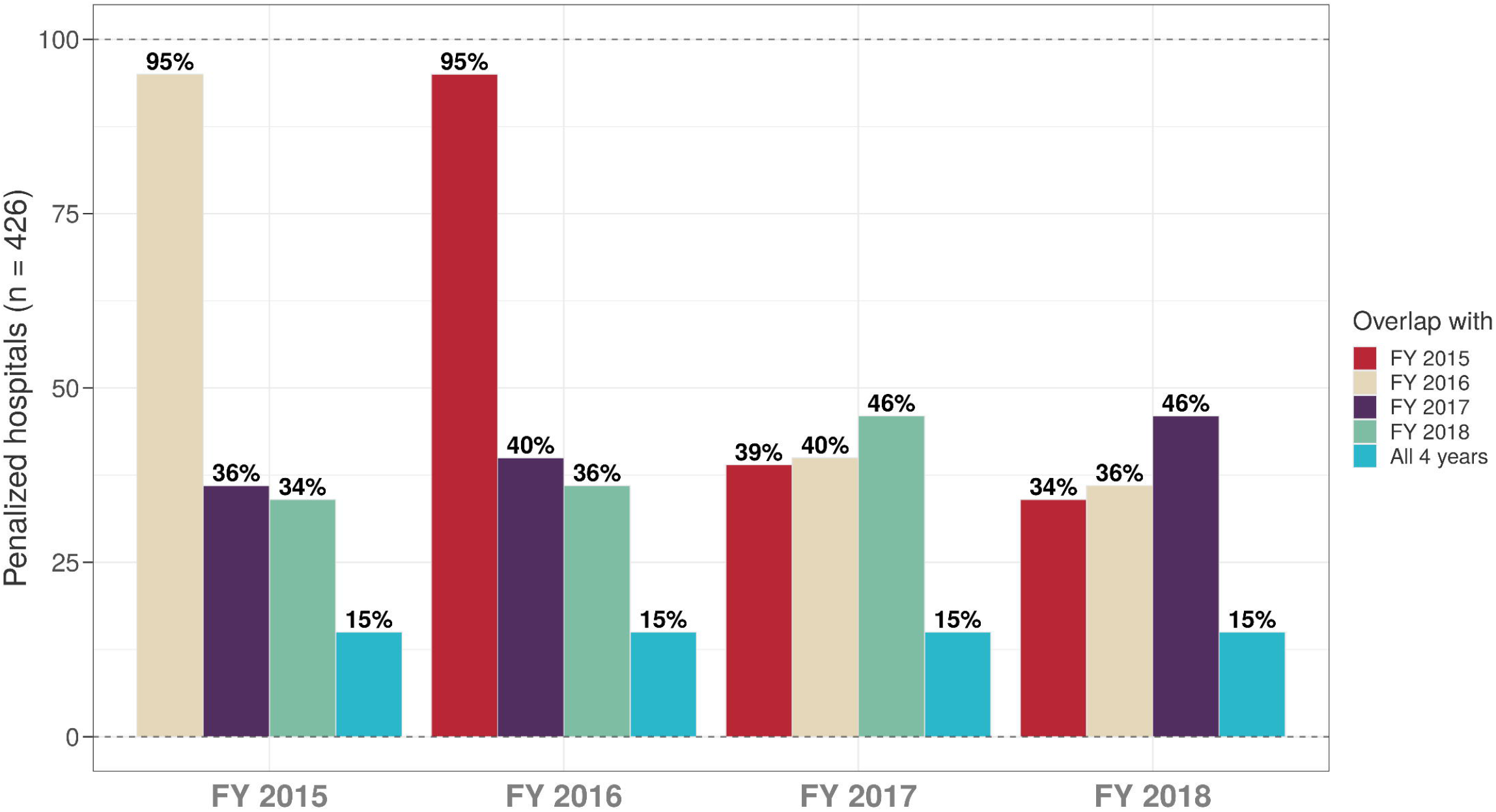
Percent overlap in penalized hospitals among FY 2015-2018 scoring methodologies

To investigate whether shifts in penalty status were due to small changes driven by tight clustering of HAC scores around the penalty threshold, we examined changes in relative rankings of all hospitals (penalized or not) based on the HACRP scoring methodology (Additional files 1: Figs. A2 and A4). We found significant changes in hospital (relative) ranking under FY 2016, 2017 and 2018 HACRP scoring methods, regardless of how Domain 2 values with missing NHSN data were assigned. These results indicate that hospital relative ranking was driven primarily by changes in Domain 1 definition.

## Discussion

In this evaluation of the Hospital-Acquired Condition Reduction Program using Medicare discharge and NHSN data for hospitals in 14 states, we report two main findings. First, we found substantial sensitivity of the HACRP penalties to annual program updates. Second, changes in hospital’s penalty status were primarily driven by changes in the definition of Domain 1.

We found the highest overlap in penalized hospitals between FY 2015-2016, reflecting relatively a modest scoring change between these two years, the introduction of SSI to Domain 2 (Exhibit 2). However, when we compared penalized hospitals between FY 2016 and 2017 scoring methods, the overlap decreased to only 40% (*n* = 169 out of 426). This decrease was completely attributable to re-weighting of individual PSIs under Domain 1, because the majority of penalized hospitals (*∼*60%) did not report any Domain 2 measures. Similarly, the lack of overlap between FY 2017 and 2018 penalized hospitals was also driven by another recalibration of PSI-90.

Our findings raise important concerns about the efficacy and fairness of the HACRP program over time. Penalty status under this program has largely been driven by recalibrations of the PSI-90, rather than any significant differences in hospital performance. Using the same performance data over four program years, we find that only 15% of hospital would have received penalties under all scoring methods. Thus, the HACRP penalties assessed (and publicly reported) are, at best, very “noisy” indicators of hospital performance, providing very little guidance to hospitals on patient safety improvement and very little information to patients seeking to identify “safer” hospitals. At worst, the HACRP program is assessing penalties on a relatively random of set of hospitals, providing additional financial resources to the Medicare program, but yielding almost no benefit to beneficiaries.

Unreliable performance measures also stimulate inefficiency in our healthcare system. Hospitals with high penalties will make investments in their quality and safety infrastructure to avoid future penalties. When these penalties reflect vagaries of measurement rather than real performance differences, many hospitals that are truly less safe will under-invest in improvement, while those that are truly safer may divert resources from other patient care improvements simply because of the penalty. The disconnect between penalties and effective hospital investments in quality improvement has been raised in the context of other Medicare penalty programs [20].

Inefficient use of resources is especially concerning for low-margin hospitals, such as those serving low-income and vulnerable populations. Safety-net and teaching hospitals are more likely to receive HACRP penalties [15]. These financial penalties, combined with an inability to effectively direct scarce resources to improve safety, may further widen healthcare disparities for populations served by these hospitals. Previous research has sounded the alarm regarding the impact of pay-for-performance penalties on health disparities [21, 22, 23]; our work adds a new facet to this debate: the detrimental impact of penalties on the ability of these important hospitals to effectively improve care for the patients they serve.

Our findings provide some insights on how to improve the HACRP program. While changes to Domain 1 and Domain 2 scoring methods were rooted in stakeholder feedback and program improvement, they effectively “moved the goalposts” after performance periods were concluded. For maximal effectiveness, changes to the program should be communicated in manner and timeframe that allows hospitals to make appropriate investments in infrastructure and changes to patient care. We would also suggest caution when considering multiple program changes over a short period of time, as these changes contribute to hospital uncertainty regarding penalty status and may limit their willingness to pursue particular improvements.

### Limitations

Due to data limitations, we were unable to include all hospitals subject to the HACRP program in our analyses. Our more limited dataset focusing on 14 states implies that the 75th percentile cutoff in our data did not necessarily match the one used by CMS to identify hospitals subject to a penalty. Because our goal was to explore the sensitivity of HACRP penalties to annual program updates, rather than reproduce CMS results, this limitation is unlikely to affect our conclusions.

Our analyses also relied on a prediction model for CLABSI and CAUTI SIRs to calculate Domain 2 measures under CMS FY 2015-2017 methodologies. This was also driven by data limitations. Nonetheless, the impact of prediction error on Domain 2 measures is likely to be minimal because FY 2015-17 penalties relied on relative ranking rather than actual values.

## Conclusion

Early evaluations of the HACRP program suggested positive impact on hospital acquired conditions. Our work, combined with earlier studies questioning the link between HACRP penalties and hospital quality of care, suggest that HACRP penalties are not reliable indicators of hospital performance, creating inefficiencies and potentially exacerbating healthcare disparities. We recommend caution when considering further modifications to the HACRP program to limit the randomness of penalties characterizing the program’s history.

## Supporting information

Supplementary results

## Data Availability

The data that support the findings of this study are available from the Healthcare Cost and Utilization Project (HCUP), but restrictions apply to the availability of these data, which were used under license for the current study, and so are not publicly available. Data are, however, available from the authors upon reasonable request and with permission of HCUP.

## Abbreviations

HACRP: Hospital acquired conditions reduction program
HAC: Hospital-acquired conditions
CMS: Centers for medicare and medicaid services
AHRQ: Agency for healthcare research and quality
CDC: Centers for disease control and prevention
PSI: Patient safety indicator
NHSN: National healthcare safety network
HAI: Healthcare associate infection
FY: fiscal year
HCUP: Healthcare cost and utilization project
SID: State inpatient databases
SIR: standardized infection ratio
CLABSI: central line-associated bloodstream infection
CAUTI: catheter-associated urinary tract infection
SSI: surgical site infection
MRSA: methicillin-resistant staphylococcus aureus
CDI: clostridium difficile infection
ICD: International classification of diseases

## Acknowledgements

Not applicable.

## Funding

This study received funding support from Agency for Healthcare Research and Quality (R01HS025148).

## Ethics approval and consent to participate

Data in this study was obtained from the Healthcare Cost and Utilization Project (HCUP) which already removed all identifying information to protect the privacy of individual patients, physicians, and hospitals.

## Competing interests

The authors declare that they have no competing interests.

## Consent for publication

Not applicable.

## Authors’ contributions

OV and TW contributed to the conception and design of the study, analysis and interpretation of data. OV, KM, PZ, and TW contributed to drafting of the manuscript and final approval of the manuscript.

## Notes

### Competing Interest Statement

The authors have declared no competing interest.

