## Supplementary results for "Measurement Matters: Changing Penalty Calculations under the Hospital Acquired Condition Reduction Program (HACRP) Cost Hospitals Millions"

**Table A1.** Descriptive statistics of total HAC scores

| | No data submitted to NHSN implies no Domain 2 score | | No data submitted to NHSN implies maximum Domain 2 score | | |
|---|---|---|---|---|---|
| | Mean (SD) | 75th percentile cutoff | Mean (SD) | 75th percentile cutoff | CMS reported 75th percentile cutoff |
| FY 2018 | -0.06 (0.72) | 0.263 | 0.94 (1.12) | 1.999 | 0.371 |
| FY 2017 | 5.72 (2.23) | 7.000 | 7.31 (2.56) | 9.400 | 6.570 |
| FY 2016 | 5.91 (2.32) | 8.000 | 7.48 (2.35) | 9.250 | 6.750 |
| FY 2015 | 5.85 (2.38) | 8.000 | 7.25 (2.31) | 8.950 | 7.000 |

Source: Author's analysis of HCUP administrative data and NHSN HAIs for hospitals in 14 states.

**Figure A2.** Changes in total HAC scores by FY-specific methodology.

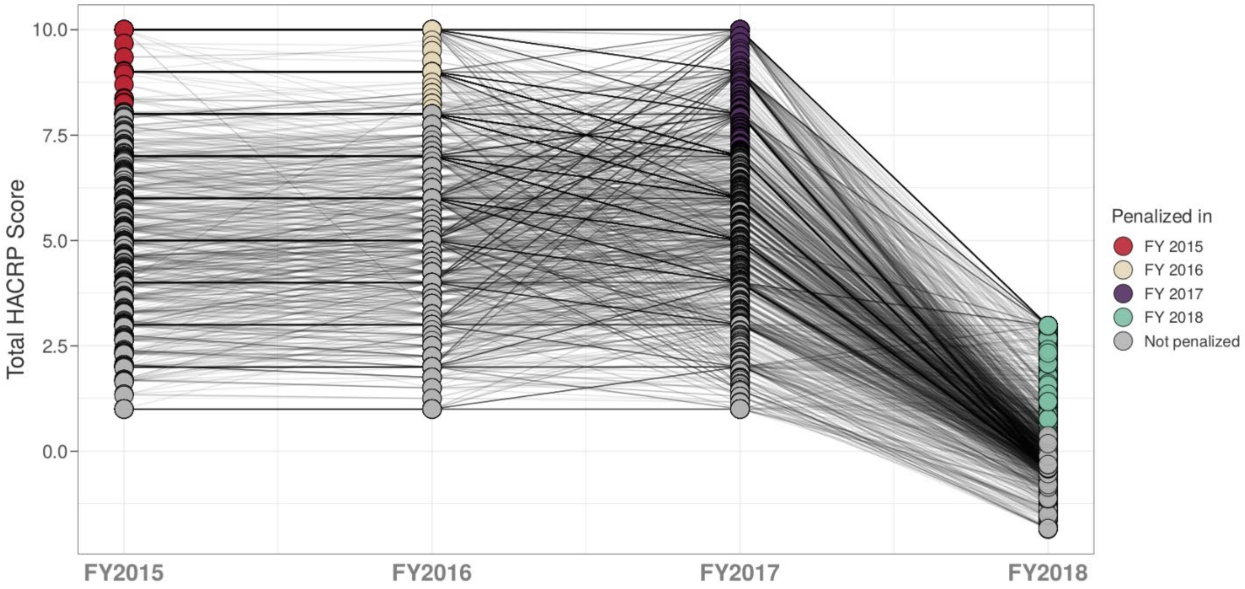


Source: Author's analysis under the assumption that if no HAI data was submitted to NHSN then no Domain 2 score.

Notes: Points in color represent total HAC scores that are greater than the 75th percentile of the empirical distribution.

**Figure A3.** Percent overlap in penalized hospitals among FY 2015-2018 scoring methodologies.

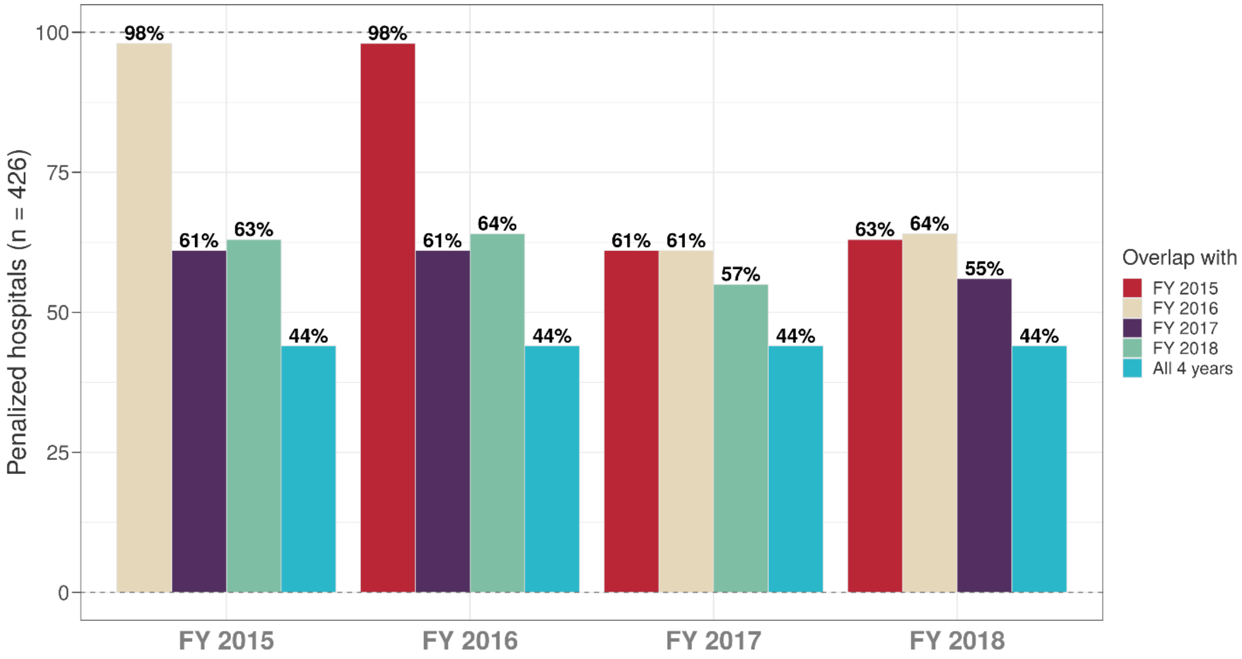


Source: Author's analysis under the assumption that if no HAI data was submitted to NHSN then maximum Domain 2 score.

**Online Exhibit A4.** Changes in total HAC scores by FY-specific methodology.

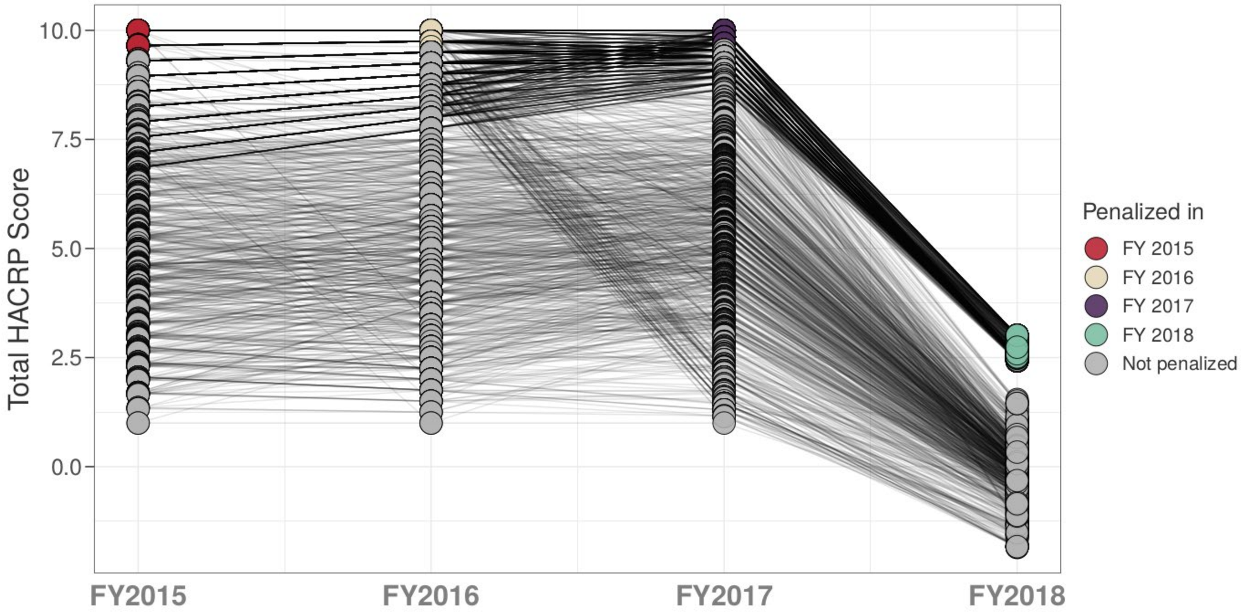


Source: Author's analysis under the assumption that if no HAI data was submitted to NHSN then maximum Domain 2 score.

Notes: Points in color represent total HACRP scores that are greater than the 75th percentile of the empirical distribution.
